# Why causal effects of ultra-processed foods cannot be identified: a systematic review of subtypes of ultra-processed foods and risk of type 2 diabetes

**DOI:** 10.1101/2025.10.14.25337888

**Authors:** Michael Fridén, Maria Mai, Anja Olsen, Christina C. Dahm, Daniel B. Ibsen

## Abstract

**Background:** One of the key assumptions underlying causal inference is the assumption of consistency; that is, the absence of relevant multiple versions of the same treatment. Although ultra-processed food (UPF) as a dietary construct has been associated with a greater risk of type 2 diabetes (T2D), subtypes of UPFs may exert different strengths and directions of associations on T2D. The main objective of this study was to systematically evaluate the current literature on subtypes of UPFs and their associations with T2D and to contextualize these findings within the framework of counterfactual theory, with particular emphasis on the consistency assumption.

**Methods:** We searched PubMed and Embase for relevant articles. Cohort studies of adult men and women without T2D and with the exposure (subtypes of UPFs) and outcome (incident T2D) of interest were included. Two independent researchers screened the articles and evaluated risk of bias using the ROBINS-E tool. UPFs were categorized into eight subtypes. No meta-analyses were conducted. A post-hoc analysis of n=52,201 in the Danish Diet, Cancer and Health (DCH) cohort was performed to illustrate the implications of causal inconsistency. We formed a fictive food group (the terrible five) combining sugar sweetened beverages, processed meat, red meat, refined grains and vegetables and investigated the associations with T2D.

**Results:** Out of n=222 articles screened for eligibility, n=6 cohort studies with n=635,332 participants (average follow-up of 6-26.1 years) were finally included. Most studies on UPF breads and cereals; packaged snacks; and sauces, spreads and condiments showed no associations with T2D. Most studies on UPF ready-to-eat-dishes; meat-based products; and artificially and sugar-sweetened beverages were positively associated with T2D. UPF dairy products; and sweets, snacks and confectionaries were generally inversely associated with T2D, although with some variation. Overall risk of bias was considered moderate to high in all included studies. In the DCH cohort, a positive association between the “terrible five” and T2D was observed (HR: 1.14, 95% CI: 1.05, 1.23). Intake of vegetables was inversely associated with T2D (HR: 0.96, 95% CI: 0.95, 0.98).

**Conclusion:** The systematic review showed different strengths and directions of associations on the risk of T2D across the different subtypes of UPFs. Aggregating these sources into a coarse food category such as UPF therefore violates the assumption of causal consistency and, as illustrated in our post hoc analysis, can mask individual associations. This study illustrates potential unintended consequences of broad implementation of UPF-related policies without further improvement of the definition or mechanistic understanding.

**Registration:** The protocol was registered in PROSPERO (ID: CRD420251058458).

## Background

Ultra-processed foods (UPFs) have consistently been associated with greater risk of various cardiometabolic diseases, including type 2 diabetes (T2D) [1]. According to the NOVA classification system, introduced by Monteiro et al., in 2009, and updated in 2019, UPFs are defined as “formulations made mostly or entirely from substances derived from foods and additives” encompassing items such as candy, soda, processed meats, sweetened yoghurt and certain types of bread [2,3]. As such, the NOVA system treats all UPFs as a homogeneous food category.

However, recent scientific discussions have raised concerns of whether different subtypes of UPFs may have varying effects on cardiometabolic diseases, including T2D risk; with some potentially even having beneficial effects [4,5]. From a causal inference perspective, using the counterfactual framework (i.e., the potential outcomes framework), this challenges a fundamental assumption, namely the assumption of *consistency* [6].

Formally, consistency implies that the observed outcome under treatment *a* is equal to the outcome that would have been observed had treatment *a* been assigned (i.e., Y*^a^ = Y for every unit with A = a)*. Informally, this assumption holds when causal questions are *sufficiently* well defined and no other relevant versions of the treatment exist [6,7]. When consistency is violated, for example when broader diet categories include diverse foods with distinct effects, causal estimates may become difficult to interpret. For instance, a meta-analysis found that a higher intake of total potatoes was associated with a higher risk of T2D. However, when dividing total potatoes into French fries and boiled, baked or mashed potatoes, a higher risk was only observed for French fries [8]. Such violations of consistency have implications for generalizing and transporting study findings across populations for public health decision making, especially if the assumption of treatment version irrelevance (i.e., that different versions of the treatment yield the same effect) does not hold [9].

An important, yet underexplored, aspect of UPFs is therefore the need to identify whether distinct UPF subtypes exert different effects on health outcomes such as T2D. The main objective of this study is therefore to systematically evaluate the current literature on subtypes of UPF and their associations with T2D and to contextualize these findings within the framework of causal inference theory, with particular emphasis on the consistency assumption.

## Methods

### Design

The Preferred Reporting Items for Systematic Reviews and Meta-Analyses (PRISMA) guidelines were adhered to [10] and the full study protocol was registered at PROSPERO (ID: CRD420251058458). Covidence was used to import references, manage data as well as to construct a study flow-chart of included studies. Two independent reviewers screened the studies (M.F., and M.M.) and assessed risk of bias (M.F., and D.B.I.). Any conflicts between researchers were resolved through internal discussion. Extraction of data was performed by M.F. using Excel.

### Data sources and searches

A systematic literature search was conducted in PubMed (last searched 6^th^ of June 2025) and Embase (last searched 9^th^ of June 2025). The search strategy was restricted to articles published in English and combined free-text terms, database-specific subject headings (MeSH in PubMed, Emtree in Embase), and Boolean operators. No time restriction was specified.

The search term for the PubMed database was: ((((((ultra-processed food*[Title/Abstract]) OR (ultraprocessed food*[Title/Abstract])) OR (UPF[Title/Abstract])) OR ("UPF intake"[Title/Abstract])) OR ("Food, Processed"[MeSH Terms]) OR ("NOVA classification"[Title/Abstract]))) AND (((("diabetes mellitus, type 2"[MeSH Terms]) OR ("type 2 diabetes"[Title/Abstract])) OR ("type two diabetes"[Title/Abstract])) OR ("diabetes mellitus type 2"[Title/Abstract]))) AND (english[lang]) whereas the search term for Embase was: (’ultra processed food*’:ab,ti OR ’ultraprocessed food*’:ab,ti OR ’upf intake’:ab,ti OR ’nova classification’:ab,ti) AND (’type 2 diabetes mellitus’/exp OR ’type 2 diabetes’:ab,ti OR ’type two diabetes’:ab,ti OR ’diabetes mellitus type 2’:ab,ti) AND [english]/lim.

### Eligibility criteria

Articles were included if they met the following eligibility criteria:

(i) Cohort study
(ii) Adult men and women aged ≥18 years without T2D at baseline
(iii) Assessment of subtypes of NOVA UPFs
(iv) Incident T2D diagnosis based on questionnaires, electronic health records, drug registries, laboratory blood sampling, or a combination of these sources

Exclusion criteria were:

(i) Studies involving children or adolescents
(ii) Lack of information on subtypes of UPF
(iii) Study designs such as case-control-, cross-sectional-, intervention-, ecological studies or case series

### Data extraction and risk of bias assessment

Associational measures (risk ratios (RRs) with 95% confidence intervals (CI)) estimated by the regression model specified as the final model by the authors were extracted by M.F. If the authors did not specify their final model, results from the fully adjusted model were extracted. Hazard ratios and odds ratios were interpreted as RRs. UPFs were categorized into the following eight overall subtypes: Breads and cereals; Sauces, spreads and condiments; Sweets, snacks and confectionary; Dairy products; Packages snacks; Artificially and sugar-sweetened beverages; Meat-based products; Ready-to-eat-dishes (**Supplementary Table 1**). Canhada 2023 [11] provided two estimates per dietary source of UPFs; one in the unit of 50 grams/day and one in the unit of 1 SD in grams/day. The former was extracted for the purpose of this analysis as SD units are highly population-specific.

Risk of bias was assessed using the ROBINS-E tool for non-randomized studies [12]. The ROBINS-E tool evaluates risk of bias across seven domains: confounding, measurement of the exposure, selection of participants into the study (or analysis), post-exposure interventions, missing data, measurement of the outcome and selection of the reported result. Confounders were selected with the help of subject-matter expertise and a directed acyclic graph (DAG), using the online software Dagitty (**Supplementary Figure 1**) [13,14]. Identified confounders were age, sex, physical activity, socioeconomic status (e.g., education), family history of T2D, total energy intake (or total food intake) and smoking. Sleep and previous diet were considered to have small effects on the exposure and the outcome and were therefore not included. BMI and other risk factors associated with T2D such as the amount of body fat, presence of insulin resistance and concentrations of fasting glucose were considered mediators on the causal pathway. Adjusting for mediators was considered overadjustment of the total effect [15]. For studies involving time-varying covariates influenced by past exposure (e.g., physical activity and diet), conventional regression-based methods for adjustment were deemed insufficient to fully control for confounding. Application of g-methods were judged to be necessary to account for exposure-confounder feedback [16].

### Statistical analysis

A meta-analysis of the results was deemed unfeasible to perform due to differences in units of measurement given rise to slightly different interpretations of individual estimates. We therefore focused on the qualitative direction of the estimates. R (version 4.5.0) was used to build graphs with the use of the following packages: “*meta*”, “*robvis*” and “*ggplot2*”.

### The terrible five and risk of T2D in the DCH cohort: a cohort study

In a post-hoc analysis of this systematic review, *de novo* results from an additional cohort study were included to illustrate the impact of causal inconsistency. For this analysis we investigated the association between adherence to a diet with higher intake of a fictive group of foods, here called the “terrible five”, including processed meat, sugar sweetened beverages, red meat, refined grains and vegetables. Previous cohorts have shown that a high intake of processed meat, sugar sweetened beverages, red meat and refined grains are associated with a higher risk of T2D whereas intake of vegetables is associated with a lower risk [17]. For the analysis we used the Danish Diet, Cancer and Health (DCH) cohort [18]. Dietary data were collected using a validated 192-item FFQ [19,20] and the outcome was obtained from registry linkage to the Danish National Diabetes Registry using each participant’s unique national personal identification number [21]. A Cox proportional hazards model with age as the underlying time-scale was fitted to the data to estimate HRs with corresponding 95% CIs of quartile comparisons of the association between high intake of the “terrible five” compared with low intake and risk of T2D as well as a model where all the individual foods per 50 g/day were included. The models were adjusted for age and calendar time at baseline in tertiles, as strata and for sex (men, woman), physical activity (<30 min/day, ≥30 min/day moderate to vigorous exercise), education (vocational, e.g. plumber and hairdresser; short, e.g. only primary school, high school or one or more short courses; medium, e.g. social worker or BSc degree; and long e.g. MSc or higher degree from university), smoking status (never, former, current), alcohol intake (g/day, continuous) and total energy intake (kJ/day, continuous). The main purpose of this post-hoc analysis was to examine if grouping foods with different directions of association towards health (suggested from previous literature) into the same category (such as harmful foods and vegetables) may lead to erroneous conclusions being drawn. Stata (version 18.0) was used for the post-hoc analysis.

## Results

The flowchart of the study is shown in **Figure 1**. After n=88 duplicates had been removed, n=222 titles and abstracts were screened for eligibility. Of these, n=28 full-texts were reviewed. After an additional removal of n=22 articles on the basis of having the wrong intervention, outcome or study design, for not presenting any results or for not being a full peer-reviewed publication, n=6 cohort studies were finally included. Duan 2022 [22] was excluded due to the broad categorizations of “Sweet snack pattern”, “Cold savory snack pattern” and “Warm savory snack pattern”, identified through data-driven principal component analyses.

**Figure 1.**
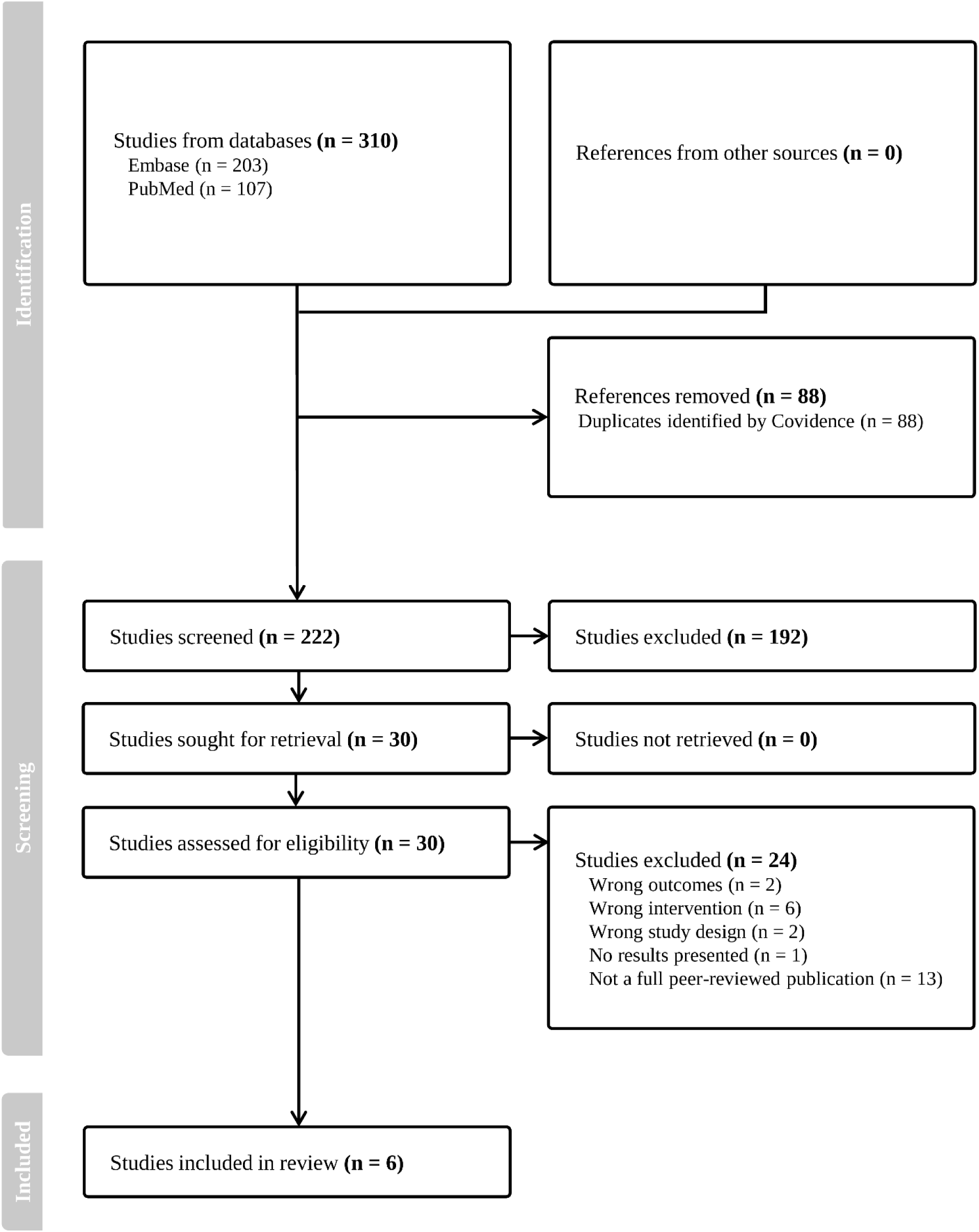
PRISMA flow-chart of included studies.

### Study characteristics

**Table 1** show the study characteristics. All included studies were published in the last 5 years and were conducted in Europe (Sweden, France, UK, Germany, Italy, Denmark, Spain and the Netherlands), North America (United States), South America (Brazil) and Asia (Iran and South Korea). The average follow-up time ranged from 6 to 26.1 years and most studies used a food-frequency questionnaire (FFQ), either alone or combined with other measurement tools, to assess habitual dietary intake at baseline. Only one study (Chen 2023) used time-updated information on subtypes of UPFs and confounders. The unit of measurement varied from grams, servings, weight ratios and energy percentages (E%) among studies. Most studies assessed T2D using a self-reported questionnaire, alone or in combination with other more objective assessment methods.

**Table 1.**
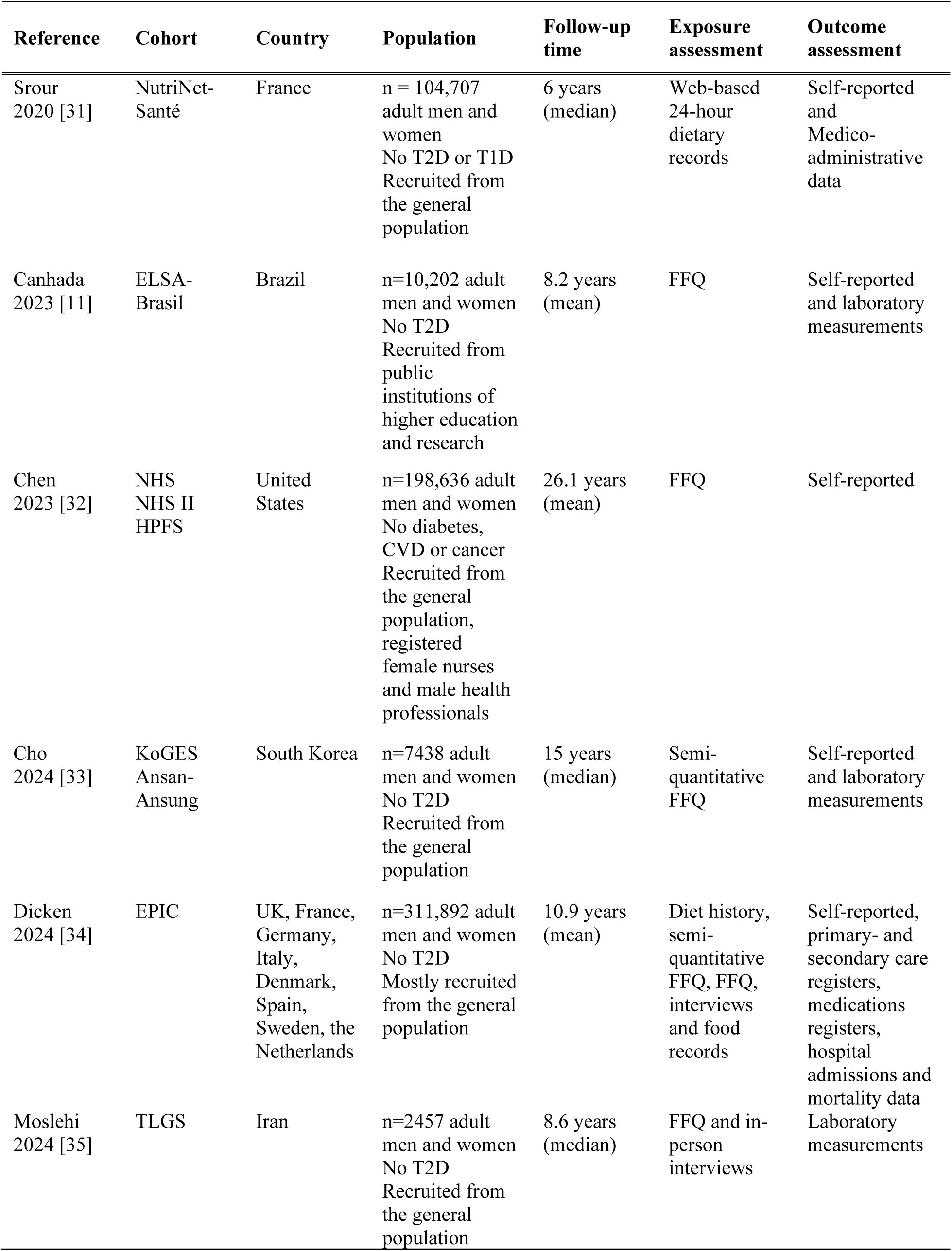
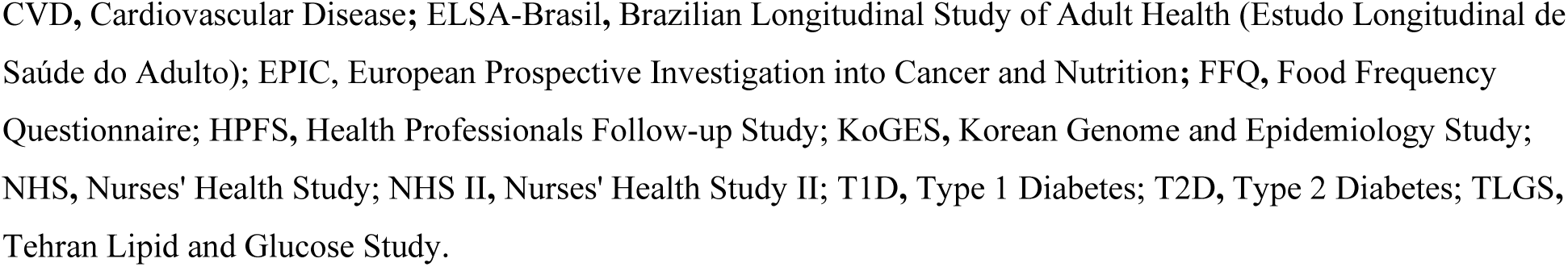
Summary characteristics of included cohort studies.

### Results from the systematic review

Results from the systematic review are shown in **Figure 2**. Most studies on UPF from breads and cereals; packaged snacks; and sauces, spreads and condiments showed no clear associations with T2D. Most studies on UPF from ready-to-eat-dishes; meat-based products; and artificially and sugar-sweetened beverages were positively associated with T2D. UPF from dairy products and sweets, snacks and confectionaries were generally inversely associated with T2D, although with some variation.

**Figure 2.**
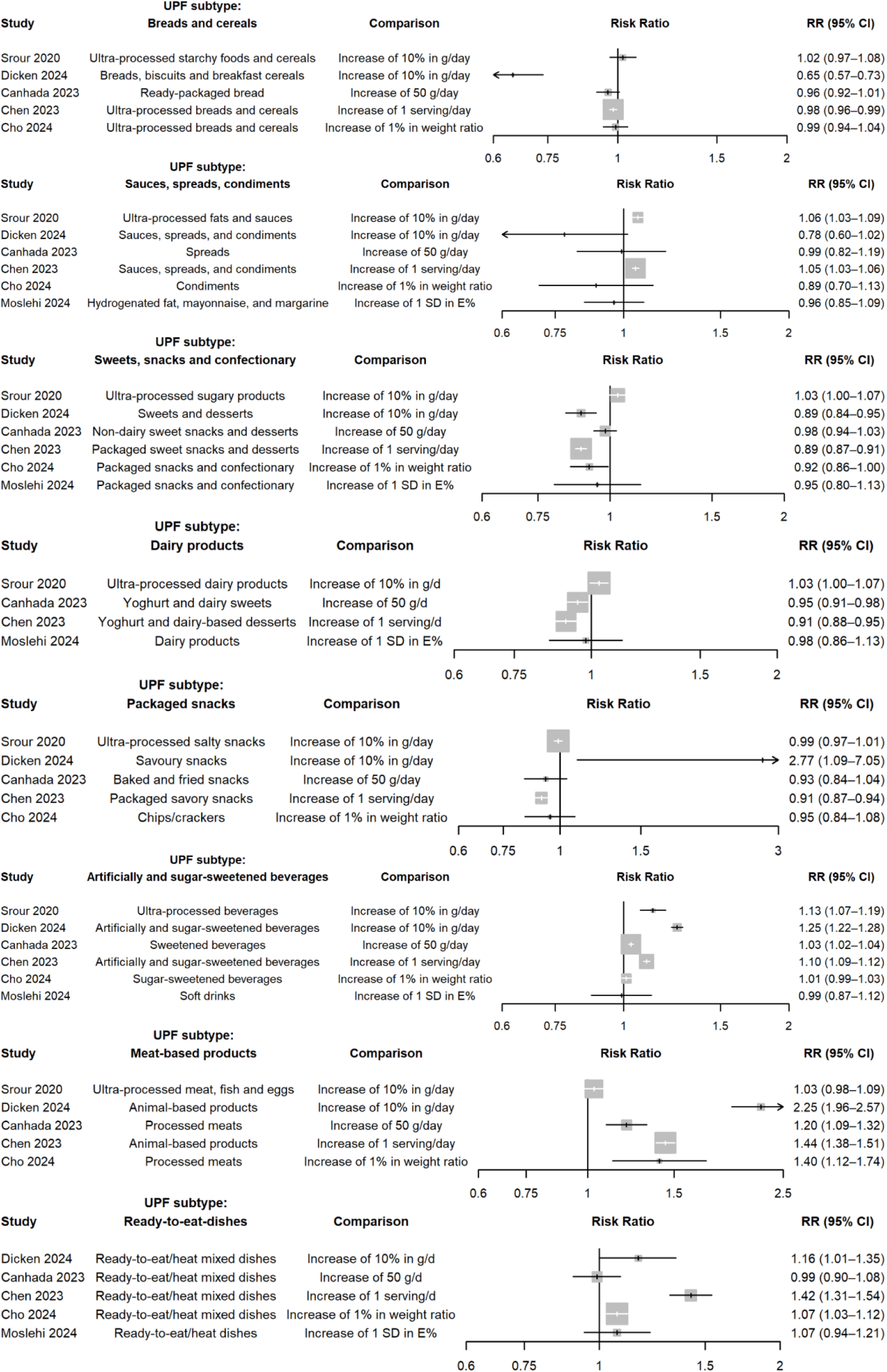
Forest plots of subtypes of UPFs and the risk of T2D.

Overall risk of bias was considered moderate to high for all included studies (**Supplementary Figure 2 and 3**). The n=4 studies that were classified with overall high risk of bias suffered from unmeasured confounding, missing data and selection bias, the latter primarily through high proportions of loss to follow-up. As no correction methods were used to adjust for loss to follow-up, such as inverse-probability weighting or multiple imputation, they were therefore considered high risk of bias. All studies used self-reported questionnaires to assess intake of UPFs and were therefore scored with “Some concerns”. All studies had low risk of bias due to selection of reported results.

### Results from the DCH cohort study

In total, n=52,201 were included in the analysis after removing those with cancer, cardiovascular disease and diabetes before baseline (n=3276), those with missing data in covariates (n= 646) and those with implausible energy intake (n=930). Characteristics of the study population are shown in **Supplementary Table 2**. After a median of 15.4 years of follow-up, a total of n=6733 developed T2D. Comparing the highest and lowest quartile, a higher intake of the “terrible five” was associated with a 14% (HR: 1.14, 95% CI: 1.05, 1.23) higher risk of T2D. In a mutually adjusted model, a 50 g/day higher intake of processed meat (HR: 1.24, 95% CI: 1.16, 1.31), sugar sweetened beverages (HR: 1.04, 95% CI: 1.03, 1.05), red meat (HR: 1.15, 95% CI: 1.12, 1.20) and refined grains (HR: 1.06, 95% CI: 1.02, 1.10) was associated with a higher risk whereas a higher intake of vegetables was associated with a lower risk (HR: 0.96, 95% CI: 0.95, 0.98) of T2D.

## Discussions

### Summary of the findings

In our systematic review of cohort studies, we found that the direction of associations between UPFs and T2D varies across the different subtypes. Most studies on breads and cereals; packaged snacks; and sauces, spreads and condiments showed no associations with T2D. Most studies on ready-to-eat-dishes; meat-based products; and artificially and sugar-sweetened beverages were positively associated with T2D. Dairy products and sweets, snacks and confectionaries were in general inversely associated with T2D, although with some variation. Overall risk of bias was considered moderate to high in all included studies.

### Strengths and limitations

Key strengths of this study include the systematic assessment of the literature using two large databases as well as the rigorous bias assessment using the ROBINS-E tool performed by two independent researchers. The rationale for not conducting a meta-analysis of the results may be viewed as a limitation. However, pooling results with different units of measurements, and thus different interpretations may not be that informative from a public health perspective, nor important for the scope of this systematic review. Similar reasoning was provided for assessing the certainty of evidence. A key limitation was that the individual studies reported slightly different definitions of the subtypes of UPF. Subtypes of UPF were grouped into six larger groups. However, if data had been more detailed and harmonized across the individual studies, further investigation of sources of heterogeneity could have been conducted. An inherent issue to all systematic reviews is the subjective assessment of risk of bias, particularly in the absence of any formal quantitative bias analyses (or other sensitivity analyses) performed by the authors. Lack of essential information on key variables (e.g., results from validation studies on self-reported outcome classifications) may also have led to errors in the bias assessment.

### Causal inconsistency – implications for future research and public health decision making

Findings from this systematic review may have important implications for guiding future research efforts. A key question in the field of UPF research is whether intake of UPF cause, for instance, T2D. When there are multiple (relevant) versions of the treatment, this causal question cannot be answered. In principle, we could have one version of a diet high in UPF including mostly processed dairy, which our included studies indicate would be associated with a lower risk and another version where the diet high in UPF includes mostly processed meat, which our included studies indicate would be associated with a higher risk of T2D. A public health consequence of this is, in an extreme case, that implementing an intervention to reduce UPF consumption would in one population lead to a lower risk whereas in another lead to a higher risk, depending on which versions were used by the public. Similar reasoning could be applied to sweets and snacks, where three out of six studies reported inverse associations with T2D risk. These somewhat unexpected findings may reflect true effects within this broad food category, bias due to misreporting, or a combination of both.

Another consequence is that potentially beneficial foods are categorized as unhealthy, when they could in fact improve health. Our *de novo* analysis illustrates this example. All current dietary guidelines recommend consumption of vegetables. However, if our “terrible five” grouping was used for policy, then we would recommend against intake of vegetables. Proposed regulatory actions, such as taxation policies, food labeling, and dietary guidelines surrounding UPF may indeed effectively target the vast majority of unhealthy foods, but risk including more nutrient-dense, health-promoting foods, such as dairy products [23]. Such propositions would likely not be endorsed for vegetables in the real-world example discussed above, and a thorough discussion is therefore needed to determine whether such measures are justified for UPF.

Effects from UPF likely arise from a complex interplay of factors, including differences in nutrient profiles (e.g., added sugar and saturated fat), additives, and structural properties [24–26]. Disentangling the mechanism(s) of any single food group is already challenging, and this complexity only increases when multiple heterogenous food groups are combined. But to move closer to a causal understanding of food processing’s impact on health, we need to understand the underlying mechanisms. For instance, a recent study investigated the role of food processing and texture, comparing diets combined of processed vs unprocessed foods and hard vs soft texture [27]. In this context, the causal question is much clearer, as a specific feature of the diet is investigated. Here, different versions of treatment may not provide particularly different results. The mechanistic knowledge would allow food manufacturers to identify suitable alternatives or reformulate existing products into healthier options [25]. The public health implications of these interventions are much more likely to be similar in different populations.

From a causal inference perspective, effect estimates from a poorly defined exposure such as UPF may, at best, be interpreted as a weighted average causal effect of all UPFs contributing to that composite food category, weighted by the distribution of those specific foods in that particular population [28]. However, even if this was a valid estimand, transporting these effect estimates to inform decisions in other populations (which is frequently done in nutrition) might be problematic if the distribution of included foods differs between the study population and the target population. Specifying the research question, and in particular, the intervention, so that no relevant multiple versions of the same treatment exist, is crucial for correct interpretation of the resulting effect estimate and for transporting findings across populations. To add further complexity, in the field of nutrition science, where the exposure of interest is most often compositional, specifying the comparator food(s) or diet is equally important, but frequently forgotten; an in-depth discussion on the concept of substitution effects has been provided previously [29,30].

### How precise is precise enough?

As discussed above, evidence from this systematic review suggests that different subtypes of UPFs may exert heterogeneous, and in some cases, opposing effects on the risk of T2D.

Importantly however, even when divided into subcategories, broad classifications such as “sauces, spreads and condiments” or “breads and cereals” remain problematic from a causal inference perspective. For instance, should we assume treatment irrelevance between butter-based sauces and tomato-based sauces without added sugars? Furthermore, does the nutritional source, e.g., butter derived from grass-fed versus conventionally raised cattle affect the potential causal effect? What about timing of consumption, context (e.g., eating alone or in a group), or preparation method? These are not trivial considerations and point to the difficulty of satisfying the consistency assumption in nutritional epidemiology.

In contrast to e.g., pharmacological exposures, where treatments such as metformin are typically administered in standard doses and formulations, dietary exposures measured, most often, via FFQs often rely on coarse categorizations. These coarse categorizations are indeed necessary from a practical and logistical point of view but may introduce substantial ambiguity and potential violations of consistency. However, whether one deems an exposure well-defined or not for a specific research question is, in the end, highly dependent on the context of the study and current scientific consensus. The consistency assumption implies that exposures must be *sufficiently* well defined [6,7]. The heart of the matter lies in the word *sufficiently*. Maybe the timing of grass-fed butter-based sauces doesn’t matter if all participants are advised to consume the sauces for breakfast between 8 and 9 am, but does it matter if the advice would be to consume it anytime between 8 am and 11 pm? Does it matter if we separate butter-based sauces rich in saturated fat from tomato-based sauces if the outcome is not influenced by the content of saturated fat but of the content of micronutrients?

The field of nutrition must reckon with the trade-off between pragmatic exposure measurements and categorizations (such as those derived from FFQs) and the need for well-defined interventions. While it may not be feasible to capture every contextual nuance, researchers should critically assess whether the exposure is defined sufficiently to meet the assumption of consistency. Greater attention to this assumption is highly warranted in nutritional epidemiology, especially when studying complex, heterogeneous exposures such as UPFs, where different foods may not only differ in magnitude of effect, but also in direction. For dietary pattern scores derived from various foods with the same direction and comparable magnitudes of effects, treatment irrelevance violations may be less of a concern (especially if clearly defined intake ranges are specified for each food), although important to bear in mind.

The point of this paper is not to dismiss the value of UPF research to date; it has undeniably advanced our general understanding and sparked important discussions, but rather to emphasize that, if we want to move beyond associational UPF–disease cohort studies and towards a deeper causal understanding of ultra-processing itself, we must recognize the current limitations of the field, whereby causal inconsistency is one of the more important ones. One proposing way forward is to put more time and effort into investigating the specific mechanisms by which different processing techniques (not just “ultra-processing” as a broad category) influence short-term intermediate outcomes in tightly controlled randomized trials. Another approach, preferably combined with mechanistic research, is to refine observational research by improving dietary assessment tools to better capture the degree of food processing. These more targeted dietary assessment tools would then enable the use of methods such as food substitution models, contrasting otherwise similar foods that differ primarily in processing degree.

## Conclusion

The systematic review showed different strengths and directions of associations on the risk of T2D across the different subtypes of UPFs. Whereas most studies on breads and cereals; packaged snacks; and sauces, spreads and condiments showed no associations with T2D, most studies on ready-to-eat-dishes; meat-based products; and artificially and sugar-sweetened beverages were positively associated with T2D. Dairy products and sweets, snacks and confectionaries, on the other hand, were generally inversely associated with T2D. Aggregating these foods into a coarse food category such as UPF therefore violates the assumption of causal consistency and, as illustrated in our post hoc analysis of the “terrible five”, can mask individual associations. This study illustrates potential unintended consequences of broad implementation of UPFs policies without further improvement of the definition or mechanistic understanding.

## Authorś contributions

M.F., M.M., and D.B.I. contributed to conception and design. M.F., and M.M. screened titles and abstracts and reviewed full-text articles. M.F., and D.B.I. assessed risk of bias. M.F. extracted the results and M.F., and M.M. wrote the first draft of the manuscript. D.B.I. conducted the analyses in the DCH cohort. All authors critically revised the manuscript and accepted the final version.

## Data availability

All data generated or analyzed for the systematic review are included in this published article (and its supplementary information files). *De novo* cohort results were derived from data from the Diet, Cancer and Health Cohort and are not publicly available due to personal information content, but can be acquired upon reasonable request from the Danish Cancer Society.

