## Supplementary Material for "Why causal effects of ultra-processed foods cannot be identified: a systematic review of subtypes of ultra-processed foods and risk of type 2 diabetes"

Michael Fridén[1,2,3], Maria Mai[4], Anja Olsen[4,5], Christina C. Dahm[4], Daniel B. Ibsen[2,4]

[1]Department of Clinical Medicine, Aarhus University, Aarhus, Denmark.

[2]Steno Diabetes Center Aarhus, Aarhus University Hospital, Aarhus, Denmark.

[3]Department of Public Health and Caring Sciences, Uppsala University, Uppsala, Sweden.

[4]Department of Public Health, Aarhus University, Aarhus, Denmark.

[5]Danish Cancer Institute, Copenhagen, Denmark.

**Supplementary Table 1**. Details on the categorization of subtypes of ultra-processed foods.

| Subtype of UPF | Srour 2020 | Dicken 2024 | Canhada 2023 | Chen 2023 | Cho 2024 | Moslehi 2024 |
|---|---|---|---|---|---|---|
| Breads and cereals | **Ultra-processed starchy foods and cereals:** NA | **Breads, biscuits and breakfast cereals:** Breads, biscuits, breakfast cereals | **Ready-packaged bread:** NA | **Ultra-processed breads and cereals:** Ultra-processed cereals, ultra-processed dark and whole-grain breads, other ultra-processed refined-grain breads | **Ultra-processed breads and cereals:** Steamed buns with red bean paste (or other fillings), other breads/pastries (cream bun, castella), loaf breads, corn flakes | |
| Sauces, spread and condiments | **Ultra-processed fats and sauces:** NA | **Sauces, spreads, and condiments:** Sauces, dressing and gravies, vegetable spread and products, margarine | **Spreads:** NA | **Sauces, spreads, and condiments:** e.g. cream cheese, ketchup, margarine, soy sauce, spread butter | **Condiments:** Coffee creamer | **Hydrogenated fat, mayonnaise, and margarine:** NA |
| Sweets, snacks and confectionary | **Ultra-processed sugary products:** NA | **Sweets and desserts:** Pastries, buns, cakes, ice cream, ice pops, frozen yoghurt, sweet snacks, industrial desserts, dairy desserts and drinks | **Non-dairy sweet snacks and desserts:** NA | **Packaged sweet snacks and desserts:** Confectionaries (e.g. chocolate bars), packaged sweet snacks (e.g. energy bars) and fruit-based products (e.g. applesauce) | **Packaged snacks and confectionary:** Chips/crackers, candy/chocolate, chocopie/cake | **Packaged snacks and confectionary:** NA |
| Dairy products | **Ultra-processed dairy products:** NA | | **Yoghurt and dairy sweets:** NA | **Yoghurt and dairy-based desserts:** e.g. ice cream, frozen yoghurt, artificially | | **Dairy products:** NA |

| | | | | | | |
|---|---|---|---|---|---|---|
| | | | | sweetened yoghurt | | |
| Packaged snacks | **Ultra-processed salty snacks:** NA | **Savoury snacks:** Packaged salty snacks | **Baked and fried snacks:** NA | **Packaged savory snacks:** Fat-free popcorn, fat-free light crackers, regular crackers | **Chips/crackers:** Chips/crackers | |
| Artificially and sugar-sweetened beverages | **Ultra-processed beverages:** NA | **Artificially and sugar-sweetened beverages:** Soft drinks, fruit drinks, iced tea, other sweetened beverages | **Sweetened beverages:** NA | **Artificially and sugar-sweetened beverages:** Artificially sweetened beverages (e.g. noncarbonated low-calorie soda), sugar-sweetened beverages (e.g. 7-up) | **Sugar-sweetened beverages:** Carbonated beverages (coke, sprite), other beverages (rice punch, yuja citron tea, etc.) | **Soft drinks:** NA |
| Meat-based products | **Ultra-processed meat, fish and eggs:** NA | **Animal-based products:** Processed meat, processed cheese | **Processed meats:** NA | **Animal-based products:** e.g. bacon, beef, pork hotdogs and processed meats | **Processed meats:** Ham/sausage, fish cake/crab stick | |
| Ready-to-eat-dishes | | **Ready-to-eat/heat mixed dishes:** Potato products, pizza and focaccia, pasta, instant and canned soups, ready meals, vegetables and legumes in ultra-processed medium, rice-based dishes | **Ready-to-eat/heat mixed dishes:** NA | **Ready-to-eat/heat mixed dishes:** Chowder or cream soup, French fries potatoes, pizza, ready-made soup from cans, soup made with bouillon | **Ready-to-eat/heat mixed dishes:** Instant noodles (ramyeon), pizza/hamburger | **Ready-to-eat/heat dishes:** NA |

NA refers to missing detailed information on included foods. UPF, Ultra-processed food.

**Supplementary Table 2.** Foods by quartiles of the terrible five in the DCH cohort[1].

| | Q1 (n=13 050) | Q2 (n=13 050) | Q3 (n=13 050) | Q4 (n=13 051) |
|---|---|---|---|---|
| Terrible five intake (g/day) | 147 (55-353) | 184 (79-399) | 216 (98-443) | 287 (133-604) |
| Age (years) | 57 (51–63) | 56 (51–63) | 55 (51–62) | 54 (50–62) |
| Men (%) | 27.5 | 40.9 | 51.4 | 67.0 |
| <9 years of education (%) | 19.8 | 15.0 | 11.9 | 11.4 |
| Current smoker (%) | 38.9 | 34.2 | 33.6 | 35.9 |
| Alcohol intake (g/day) | 11.2 (1.4-43.6) | 12.7 (1.9-45.2) | 14.4 (2.3-47.3) | 15.8 (2.6-53.8) |
| Body mass index (kg/m^2) | 25.2 (21.1–31.0) | 25.4 (21.4–30.9) | 25.4 (21.4-30.9) | 25.8 (21.8-31.1) |
| <30 min/day physical activity (%) | 64.2 | 62.1 | 59.2 | 56.3 |

[1]Values are presented as pseudo medians (percentiles) for continuous variables and as percentages for categorical variables. For continuous variables, if, less than 5 people have a specific value, then the average of the 5 surrounding values are used.

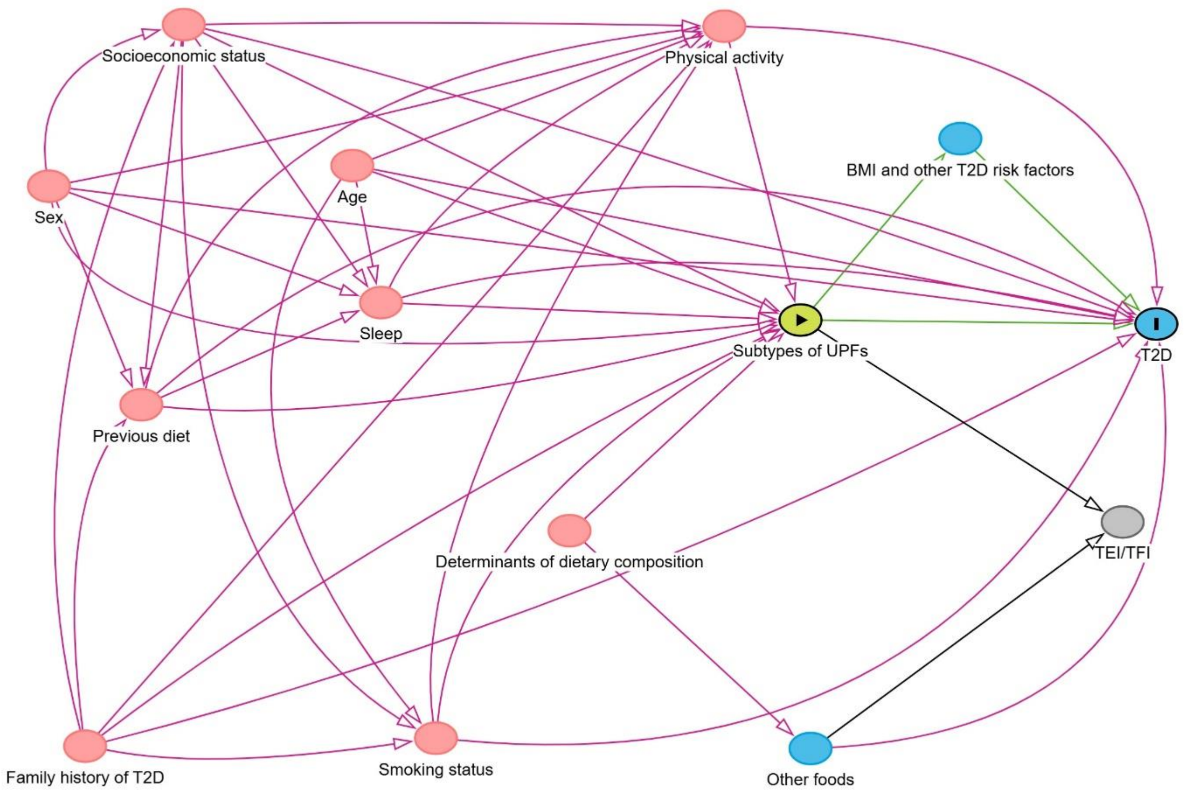


**Supplementary Figure 1**. A Directed Acyclic Graph (DAG) depicting our underlying causal assumptions between subtypes of ultra-processed foods (UPFs) and the risk of type-2 diabetes (T2D). DAGs are non-parametric graphical tools with a set of mathematical rules that are useful for identifying confounding paths (i.e., backdoor paths) to close in order to estimate unbiased causal effects. Each node in the graph connects to other nodes via arrows (or arcs). An arrow between two nodes is unidirectional (the word *directed* in DAG) and no pathway starting at one node can lead back to the same node (the word *acyclic* in DAG). Closing backdoor paths (illustrated by pink arrows in the Dagitty figure) may be achieved through various methods, such as restriction, matching and outcome regression adjustment. A mediating pathway (e.g., "Subtypes of UPFs" → "BMI and other T2D risk factors" → "T2D") is an open causal pathway that should not be conditioned on if the total effect estimand (i.e., the combined effects from all open causal pathways between the exposure and the outcome) is sought. Furthermore, conditioning on a mediator may induce collider bias if any unmeasured confounders between the mediator and the outcome are present. Total energy intake (TEI) or total food intake (TFI) act as proxy variables for other determinants of dietary composition that are commonly unobserved (such as metabolic efficiency and body size) or unknown. Sleep and previous diet were considered to have small effects on the exposure and the outcome and were therefore not included in the final adjustment set.

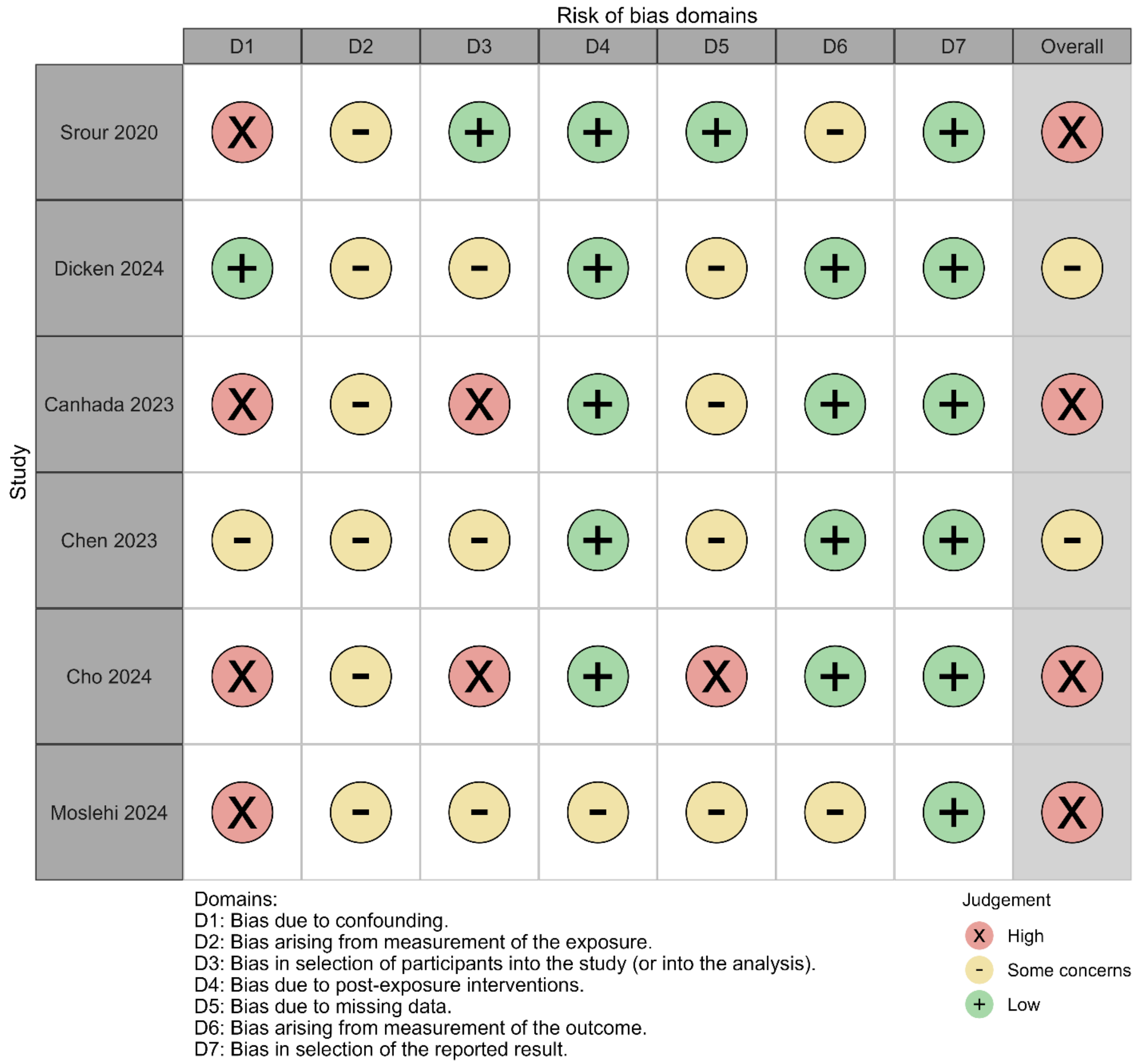


**Supplementary Figure 2**. Risk of bias assessment using the ROBINS-E tool for each individual study.

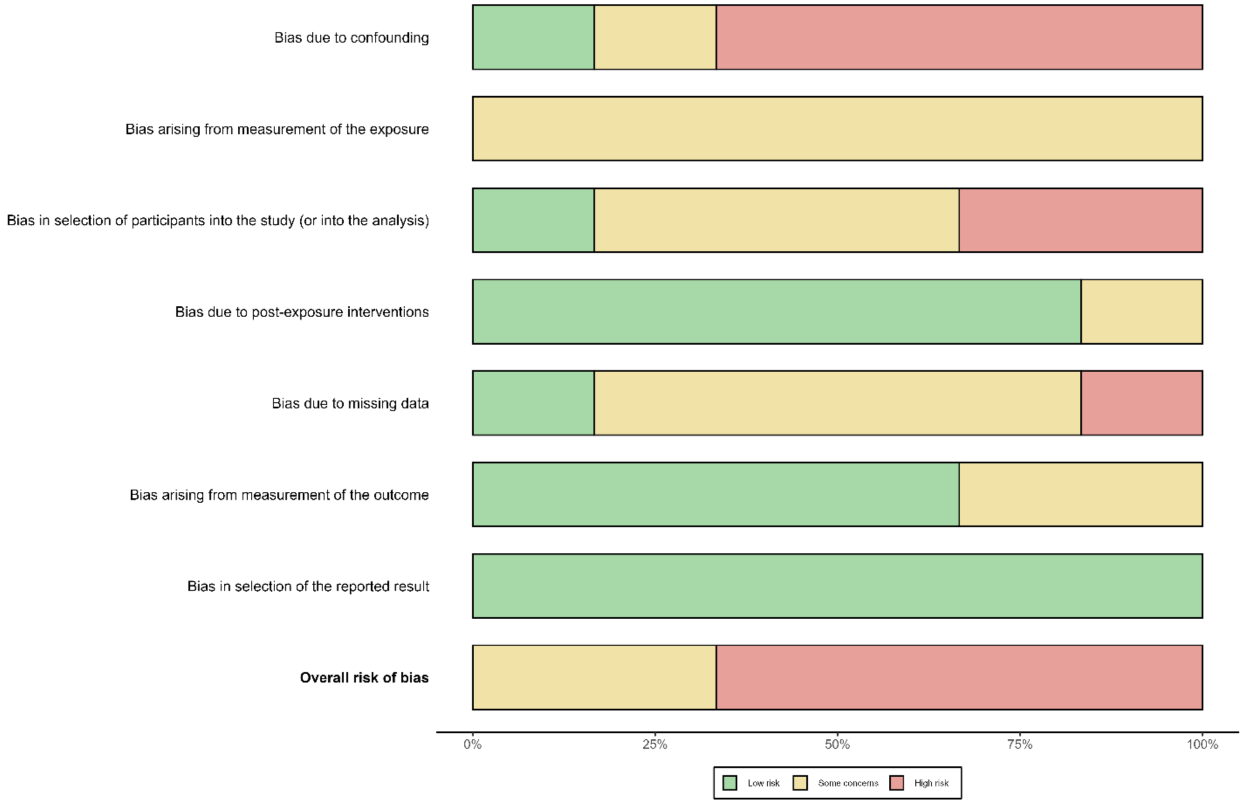


**Supplementary Figure 3**. Summary plot for the risk of bias assessment using the ROBINS-E tool.
